# Functional immunoparalysis characterized by elevated Interleukin-10 and Interleukin-10- to-Lymphocyte Count Ratio is associated with severe disease and poor outcomes in coronavirus disease 2019 (COVID-19)

**DOI:** 10.1101/2020.09.28.20203398

**Authors:** Brandon Michael Henry, Stefanie W. Benoit, Jens Vikse, Brandon A. Berger, Christina Pulvino, Jonathan Hoehn, James Rose, Maria Helena Santos de Oliveira, Giuseppe Lippi, Justin L. Benoit

**Author notes:** Correspondence to: Brandon Michael Henry, MD, Cincinnati Children’s Hospital Medical Center 3333 Burnet Ave., Cincinnati, OH, USA 45229, /.

## Abstract

**Objectives:** Severe coronavirus disease 2019 (COVID-19) is associated with a dysregulated immune state, called cytokine storm. While research has focused on the hyperinflammation, little research has been performed on the compensatory anti-inflammatory response which if severe may lead to a state of functional immunoparalysis. The aim of this study was to evaluate the anti-inflammatory response to COVID-19, by assessing interleukin-10 (IL-10) and IL-10/lymphocyte count ratio and their association with patient outcomes.

**Methods:** Adult patients presenting to the emergency department (ED) with laboratory-confirmed COVID-19 were recruited. The primary endpoint was peak COVID-19 severity within 30 days of index ED visit. Additional endpoints included COVID-19 severity at ED disposition, development of severe acute kidney injury (AKI) or secondary bacterial infections.

**Results:** A total of 52 COVID-19 patients were enrolled. IL-10 and IL-10/lymphocyte count were significantly higher in patients with severe disease at both time points (all p<0.05), as well as in those who developed severe AKI and secondary bacterial infection (all p≤0.01). In multivariable analysis, a one-unit increase in IL-10 was associated with 42% increased odds of severe COVID-19 (p=0.031), whilst a one-unit increase IL-10/lymphocyte ratio was also associated with 32% increase in odds of severe COVID-19 (p=0.013).

**Conclusions:** The hyperinflammatory response to COVID-19 is accompanied by a simultaneous anti-inflammatory response, which is associated with poor outcomes and may increase the risk of secondary bacterial infections. IL-10 and IL-10/lymphocyte ratio at ED presentation were independent predictors of COVID-19 severity. Functional immunoparalysis in COVID-19 requires further investigation to enable more precise immunomodulatory therapy against SARS-CoV-2.

## 1. INTRODUCTION

Coronavirus disease 2019 (COVID-19) is caused by the severe acute respiratory syndrome coronavirus-2 (SARS-CoV-2). Early reports of severe COVID-19 cases described a constellation of hyperinflammation and hypercoagulation, culminating in life-threatening immune-mediated and thrombotic organ damage. Excessive production of pro-inflammatory cytokines (frequently referred to as “cytokine storm”) is considered a central part of the immunopathology in COVID-19 [1], and major efforts have been made to decipher the hyperinflammatory immunophenotype and employ efficient and targeted immunosuppressive treatment strategies. Many authors have argued for the use of tocilizumab, a monoclonal anti-interleukin (IL)-6 receptor antibody, in severe COVID-19 [2]. However, in the recent randomized, double-blind, placebo-controlled phase III COVACTA trial, this drug failed to reach its primary and key secondary endpoints of improved clinical status and reduced mortality, respectively [3]. This evidence is hence reflective of a much more complex immunopathology.

Hyperinflammation may coexist with a compensatory anti-inflammatory response syndrome (CARS) in sepsis and critical illness, which induces quantitative and qualitative defects in the innate and adaptive immune system [4,5]. When severe and persistent, CARS is referred to as “immunoparalysis”, and has been associated with impaired anti-microbial defense, risk of superinfections and increased mortality [6,7]. The immunophenotype of CARS includes increased production of anti-inflammatory cytokines such as IL-10, inefficient antigen presentation, lymphocyte apoptosis and upregulation of inhibitory molecules such as programmed cell death protein (PD)-1 [4]. Molecular analyses in CARS typically reveal downregulation of Human Leukocyte Antigen – DR isotype (HLA-DR) on monocytes, as well as impaired lipopolysaccharide (LPS)-induced tumor necrosis factor (TNF)-α release ex vivo [4].

The relative presence of hyperinflammation and immunoparalysis in COVID-19 is likely to vary among patients and during an individual’s course of disease. Safe and efficient immunomodulatory therapy must be tailored to the underlying immunophenotype for preventing iatrogenic deterioration of immune function. Notably, other abnormalities may exacerbate such a functional immunoparalysis in COVID-19. A kaleidoscope of profound blood cells abnormalities have been evidenced in patients with COVID-19, encompassing especially altered myelopoiesis with generation of immature and dysfunctional leukocytes, which may hence be ineffective to counteract secondary bacterial infections [8]. Efforts should hence be made to better characterize the immunobiology of COVID-19 and develop strategies that allow for immunophenotypic stratification of patients.

In this study, we aim to characterize the anti-inflammatory cytokine profile, with emphasis on IL-10 and the IL-10/lymphocyte count ratio, in adult patients with laboratory confirmed COVID-19, and associate evidence of CARS with presence of hyperinflammation and bacterial superinfection, as well as organ damage and disease severity.

## 2. METHODS

### 2.1 Study Design

This study was performed on the Cincinnati Emergency Department (ED) COVID-19 cohort. Adults (≥18 years) who presented to the ED of the University of Cincinnati Medical Center (UCMC) between April-May 2020 with COVID-19 symptoms and underwent a clinically-necessary blood draw were preliminarily recruited in this prospective, observational study. Only patients with positive result on standard of care reverse transcriptase polymerase chain reaction (RT-PCR) test for SARS-CoV-2 in nasopharyngeal swab could be included in the final cohort. Patients with negative RT-PCR, or <18 years of age at time of presentation, or known prisoners, were excluded. This study was approved by the Institutional Review Board of the University of Cincinnati and performed under a waiver of informed consent. This study was performed in agreement with the Declaration of Helsinki and in compliance with local and national regulations.

### 2.2 Sample Collection and Processing

Blood samples were collected at index ED visit during a clinically routine blood draw. Samples were centrifuged at 2,000 g for 15 min at 4°C and subsequently frozen at -80°C until measurement. All laboratory analyses were performed at the Clinical Nephrology Lab of the Cincinnati Children’s Hospital Medical Center, with exception of routine complete blood cell counts (CBC) and serum creatinine, which were performed at the central laboratory of the UCMC. The CBC with differential was performed using a Beckman Coulter UniCel DxH 800 Cellular Analysis System (Brea, California, USA). Serum creatinine was measured using a kinetic alkaline picrate (modified Jaffe) method on either a Beckman Coulter AU480 Chemistry Analyzer (Brea, California, USA) or a Beckman Coulter AU5822 Chemistry Analyzer (Brea, California, USA).

### 2.3 Measurement of Cytokines and Other Biomarkers

Plasma concentrations of interferon (IFN)-α2a, IFN-γ, interleukin (IL)-1β, IL-6, IL-8, IL-10, IL-1 receptor antagonist (IL-1RA), and tumor necrosis factor-α (TNF-α) were quantified using Meso Scale Discovery (MSD) U-Plex assay (Rockville, Maryland, USA). Monocyte chemoattractant protein-1 (MCP-1) (R&D Systems, Minneapolis, MN, USA) and CD163 (Diapharma Group, Inc., West Chester, OH, USA) were assayed using an enzyme linked immunosorbent assays (ELISA). Plasma concentrations of fibrinogen, ferritin, myoglobin, haptoglobin, and C-reactive protein were measured using a BN II System (Siemens Medical Solutions USA, Inc., Malvern, PA, USA). Lactate dehydrogenase was measured on Dimension RxL Max Integrated Chemistry System (Siemens Medical Solutions USA, Inc, Malvern, PA, USA), while procalcitonin was tested with a chemiluminescent immunoassay (CLIA) on the Diasorin Liaison XL (DiaSorin S.p.A. Saluggia, Italy). All assays were performed according to manufacturers’ instructors and recommendations.

### 2.4 Data Collection

Data on patient demographics, past medical history, presenting vital signs, and clinical course were extracted from electronic medical records (EMR) and recorded into a REDCap (Research Electronic Data Capture) database. An ED physician performed the data extraction, with select records checked by a second ED physician for accuracy. Data on the clinical outcomes of hospitalized patients was recorded until discharge, while information on clinical course of patients discharged at index ED visit were monitored for 30 days.

### 2.5 Outcomes

The primary cytokines of interest for evaluating the compensatory anti-inflammatory response to COVID-19 were IL-10 and the ratios IL-10/lymphocyte count and IL-10/TNF-α, which have been previously used as markers of immunoparalysis [9,10]. The primary outcome was peak severity during hospitalization or within 30 days of index ED presentation. Patient severity was first quantified using a modified version of 8-point ordinal scale as outlined by the World Health Organization (WHO) R&D Blueprint [11]: (i) not hospitalized/ambulatory, no limitations of activities; (ii) not hospitalized/ambulatory, limitation of activities; (iii) hospitalized, not requiring oxygen therapy; (iv) hospitalized, requiring supplemental oxygen by mask or nasal cannula; (v) hospitalized, on non-invasive ventilation or high flow oxygen devices, or requiring intensive care unit admission; (vi) hospitalized, on invasive mechanical ventilation; (vii) mechanically ventilated and suffering from multi-organ dysfunction syndrome requiring vasopressors or extracorporeal membrane oxygenation (ECMO) or renal replacement therapy; and (viii) death. The highest ordinal scale value was documented. Patients were then classified into one of 3 severity groups, as follows: mild (ordinal scale 1-2), moderate (ordinal scale 3-4) or severe (ordinal scale 5-8).

The secondary outcomes of this study were COVID-19 severity as described above at ED disposition, and development of severe acute kidney injury (AKI) during hospitalization defined as Kidney Disease Improving Global Outcomes (KDIGO) stages 2 and 3, based on serum creatinine criteria [12]. The tertiary outcome was positive blood or urine bacterial culture throughout the course of hospitalization.

### 2.6 Statistical Analysis

Categorical data were reported as absolute number (n) and relative frequency (%), whilst continuous variables were reported as median and interquartile range (IQR). According to expected accounts, categorical variables were compared using the chi-squared test (χ^2^) or Fisher’s exact test, as appropriate. Continuous variables were compared using the Mann-Whitney *U* test. Ratios were calculated to characterize the relationships between pro-inflammatory, anti-inflammatory, and antiviral cytokines. Associations between cytokines vales were tested with Spearman’s correlation. Multivariable logistic regression was employed to identify select anti-inflammatory laboratory variables independently predicting primary and secondary outcomes. All baseline patient characteristics from Table 1 with a p-value of <0.10 were included in the model, and relevant variable selection was performed using the stepwise algorithm. Adjusted odds ratios (ORs) and 95% confidence intervals (95% CIs) were calculated. Statistical analysis was performed using Prism 8 (GraphPad Software, San Diego, CA, USA) and R (version 4.0.2, R Foundation for Statistical Computing, Vienna, Austria) with a p<0.05 considered statistically significant.

**Table 1.** Baseline characteristics of COVID-19 positive patients.

| Variable | All Patients<br>(n=52) | Maximum 30-Day Severity |  |  |
| --- | --- | --- | --- | --- |
|  |  | Mild/Moderate<br>(n=36) | Severe<br>(n=16) | p-value |
| **Age (years): median (IQR)** | 51 (39-66) | 46 (36-59) | 66 (53-71) | 0.004 |
| **Sex (male): n (%)** | 30 (57.7%) | 20 (55.5%) | 10 (62.5%) | 0.639 |
| **Comorbidities: n (%)** |  |  |  |  |
| Coronary Artery Disease | 8 (15.4%) | 5 (13.9%) | 3 (18.8%) | 0.689 |
| Heart Failure | 9 (17.3%) | 4 (11.1%) | 5 (31.3%) | 0.113 |
| Hypertension | 26 (50.0%) | 15 (41.7%) | 11 (68.8%) | 0.071 |
| Hyperlipidemia | 15 (28.8%) | 9 (25.0%) | 6 (37.5%) | 0.358 |
| Diabetes | 21 (40.4%) | 16 (44.4%) | 5 (31.3%) | 0.370 |
| Chronic Obstructive Pulmonary Disease | 8 (15.4%) | 2 (5.5%) | 6 (37.5%) | 0.007 |
| Asthma | 8 (15.4%) | 6 (16.7%) | 2 (12.5%) | 1.000 |
| Chronic Kidney Disease | 6 (11.5%) | 3 (8.3%) | 3 (18.8%) | 0.356 |
| Chronic Liver Disease | 7 (13.5%) | 3 (8.3%) | 4 (25.0%) | 0.182 |
| Cerebrovascular Disease | 7 (13.5%) | 3 (8.3%) | 4 (25.0%) | 0.182 |
| Cancer | 4 (8.0%) | 1 (2.8%) | 3 (18.8%) | 0.081 |
| Obesity | 16 (30.8%) | 11 (30.6%) | 5 (31.3%) | 0.960 |
| Inherited Immunodeficiency | 0 (0%) | 0 (0%) | 0 (0%) | - |
| Acquired Immunodeficiency (HIV, Transplant) | 3 (5.8%) | 1 (2.8%) | 2 (12.5%) | 0.165 |
| Autoimmune Disease | 2 (3.8%) | 1 (2.8%) | 1 (6.3%) | 0.547 |
| Current Smoker | 12 (23.1%) | 7 (19.4%) | 5 (31.3%) | 0.351 |
| Former Smoker | 11 (21.2%) | 7 (19.4%) | 4 (25.0%) | 0.650 |
| **Symptoms at Admission: n (%)** |  |  |  |  |
| Fever | 27 (51.9%) | 18 (50.0%) | 9 (56.3%) | 0.677 |
| Fatigue | 9 (17.3%) | 4 (11.1%) | 5 (31.3%) | 0.113 |
| Abdominal Pain | 4 (8.0%) | 3 (8.3%) | 1 (6.3%) | 1.000 |
| Nausea | 9 (17.3%) | 8 (22.2%) | 1 (6.3%) | 0.245 |
| Vomiting | 3 (5.8%) | 2 (5.6%) | 1 (6.3%) | 1.000 |
| Diarrhea | 7 (13.5%) | 2 (5.6%) | 5 (31.3%) | 0.023 |
| Headache | 6 (11.5%) | 6 (16.7%) | 0 (0%) | 0.160 |
| Sore Throat | 4 (8.0%) | 4 (11.1%) | 0 (0%) | 0.299 |
| Loss of Smell | 2 (3.8%) | 1 (2.8%) | 0 (0%) | 1.000 |
| Loss of Taste | 2 (3.8%) | 1 (2.8%) | 1 (6.3%) | 0.525 |
| Cough | 24 (46.2%) | 17 (47.2%) | 7 (43.8%) | 1.000 |
| Dyspnea | 32 (61.5%) | 23 (63.9%) | 9 (56.3%) | 0.759 |
| Chest Pain | 12 (23.1%) | 8 (22.2%) | 4 (25.0%) | 1.000 |
| Myalgia | 8 (15.4%) | 8 (22.2%) | 0 (0%) | 0.047 |
| **Days Since Symptom Onset: median (IQR)** | 7 (3-10) | 7 (4-9) | 5 (1-10) | 0.551 |

## RESULTS

### 3.1 Patient Cohort and Outcomes

A total of 52 patients with laboratory-confirmed COVID-19 were enrolled in the cohort, whose essential characteristics are shown in Table 1. At index ED visit, 46 (88.5%) patients had mild/moderate disease, while 6 (11.5%) presented with severe disease. Over the course of illness, 10 patients with mild/moderate disease progressed to severe disease, so that a total number of 16 (30.8%) patients reached the primary endpoint of severe COVID-19 within 30 days of index ED visit. Patients with severe disease were a median difference of 20 years older than patients with mild/moderate disease (p=0.004). The only co-morbidity associated with severe disease in the cohort was chronic obstructive pulmonary disease, which was observed in 37.5% of patients with severe disease as opposed to only 5.5% with mild disease (p=0.007). A total of 12 (23.1%) patients reached the secondary endpoint of severe AKI during hospitalization, 5 patients with stage 2 disease and 7 patients with stage 3 disease. Eight (15.4%) patients reached the tertiary outcome of secondary bacterial infection during hospitalization, 2 with moderate and 6 with severe COVID-19.

### 3.2 Inflammatory Biomarkers and Cytokines

Data on inflammatory biomarkers and cytokines stratified by severity at ED disposition and maximum 30-day severity are presented in Table 2. IL-6, IL-8, and IL-10 were significantly elevated in patients with severe disease both at ED disposition and within 30 days of index ED visit (all p<0.01). IL-1RA was only elevated in patients with severe disease at ED disposition (p=0.048), while TNF-α was marginally elevated in those who developed severe disease within 30 days of their index ED visit (p=0.049). To compare the relationship between pro- and anti-inflammatory responses in COVID-19, we correlated IL-6 with IL-10, and a significant positive correlation was observed (r=0.748; p<0.001).

**Table 2.** Inflammatory and Cytokine Profile of Mild/Moderate versus Severe COVID-19 patients.

| Lab Variable<br>(unit) | ED Disposition Severity |  |  | Maximum 30-day Severity |  |  |
| --- | --- | --- | --- | --- | --- | --- |
|  | Mild/Moderate<br>(n=46) | Severe<br>(n=6) | p-value | Mild/Moderate<br>(n=36) | Severe<br>(n=16) | p-value |
| White Blood Cell Count ($\times 10^9/L$) | 6.3 (5.3-9.3) | 9.6 (4.8-15.9) | 0.319 | 6.7 (5.2-9.3) | 6.8 (5.3-9.6) | 0.862 |
| Neutrophil Count ($\times 10^9/L$) | 4.4 (3.5-6.7) | 8.2 (3.4-12.9) | 0.151 | 5.2 (3.9-8.8) | 4.4 (3.3-6.7) | 0.212 |
| Lymphocyte Count ($\times 10^9/L$) | 1.0 (0.7-1.4) | 0.9 (0.4-1.5) | 0.483 | 1.0 (0.8-1.6) | 0.8 (0.6-1.2) | 0.143 |
| Platelet Count ($\times 10^9/L$) | 206 (164-263) | 220 (134-270) | 0.994 | 203.5 (163.3-252.0) | 211.5 (152.5-346.8) | 0.484 |
| Neutrophil-to-Lymphocyte Ratio | 5.3 (2.8-7.0) | 8.7 (5.6-16.8) | 0.056 | 5.3 (2.7-6.7) | 6.8 (4.3-9.7) | 0.176 |
| Platelet-to-Lymphocyte Ratio | 183.7 (134.2-296.4) | 235.8 (154.4-485.9) | 0.523 | 181.2 (118.2-267.5) | 256.4 (169.8-415.1) | 0.048 |
| C-reactive protein (mg/L) | 48.0 (8.0-121.3) | 75.0 (16.0-139.0) | 0.456 | 48.0 (7.0-122.0) | 49.0 (27.0-123.0) | 0.223 |
| Procalcitonin ($\mu g/L$) | 0.08 (0.05-0.28) | 0.32 (0.15-3.03) | 0.044 | 0.07 (0.05-0.20) | 0.26 (0.11-0.77) | 0.026 |
| Ferritin (ug/L) | 331.5 (112.8-1288.0) | 584.5 (344.3-1943.0) | 0.251 | 251.5 (106.0-1018.0) | 1032.0 (232.5-1475.0) | 0.019 |
| Lactate Dehydrogenase (U/L) | 307.5 (243.8-453.0) | 331.5 (254.8-506.0) | 0.809 | 280.0 (225.8-382.5) | 407.0 (311.0-576.0) | 0.003 |
| Haptoglobin (mg/L) | 2540.0 (1585.0-3323.0) | 2535.0 (2195.0-4313.0) | 0.454 | 2445.0 (1575.0-3315.0) | 2690.0 (2155.0-3778.0) | 0.378 |
| Myoglobin | 28.4 (17.4-96.8) | 242.5 (83.5- | 0.047 | 24.9 (15.3-65.2) | 108.0 (34.9- | 0.009 |
| (µg/L) |  | 367.0) |  |  | 342.9) |  |
| **IFN-α2a (ng/L)** | 13.1 (3.4-30.6) | 18.9 (4.9-28.6) | 0.837 | 13.3 (3.7-25.0) | 15.6 (4.6-40.7) | 0.797 |
| **IFN-γ (ng/L)** | 73.6 (40.9-279.5) | 158.1 (20.2-655.0) | 0.738 | 67.7 (30.0-276.9) | 193.5 (40.9-806.6) | 0.260 |
| **IL-1β (ng/L)** | 0.18 (0.06-0.42) | 0.78 (0.16-2.1) | 0.138 | 0.16 (0.05-0.35) | 0.36 (0.16-0.77) | 0.071 |
| **IL-6 (ng/L)** | 14.7 (4.9-33.3) | 63.2 (30.1-153.6) | 0.004 | 9.7 (2.3-19.0) | 36.4 (16.3-49.4) | <0.001 |
| **IL-8 (ng/L)** | 12.6 (7.9-23.5) | 73.8 (16.0-93.0) | 0.005 | 11.4 (5.4-17.4) | 26.8 (17.4-56.6) | <0.001 |
| **IL-10 (ng/L)** | 1.2 (0.5-2.5) | 5.9 (5.0-29.8) | <0.001 | 0.9 (0.3-2.2) | 5.3 (1.8-6.4) | <0.001 |
| **IL-1RA (ng/L)** | 1024.0 (443.6-2530.0) | 2140.0 (1518.0-3860.0) | 0.049 | 1024.0 (421.5-1908.0) | 2055.0 (842.2-3012.0) | 0.083 |
| **TNF-α (ng/L)** | 2.7 (2.0-5.1) | 8.8 (2.5-12.1) | 0.121 | 2.6 (2.1-4.0) | 5.2 (2.1-11.9) | 0.049 |
| **CD163 (ug/L)** | 1075.0 (3.2-1701.0) | 845.5 (5.3-1707.0) | 0.685 | 1055.0 (2.9-1555.0) | 1130.0 (118.2-2567.0) | 0.473 |
| **MCP-1 (ng/L)** | 530.6 (384.8-795.9) | 778.6 (556.2-1727.0) | 0.121 | 501.6 (357.4-752.8) | 713.1 (467.4-1009.0) | 0.057 |

\* All data presented as median (IQR). \*\*IL – interleukin, TNF – Tumor Necrosis Factor

Data on cytokine ratios evaluating CARS (immune dysfunction) by severity at ED disposition and maximum 30-day severity are presented in Table 3. Two markers of CARS, IL-10/TNF-α and IL-10/lymphocyte count, were significantly elevated in patients with severe disease at both time points (all p<0.05). The IL-6/lymphocyte count ratio was significantly elevated in patients with severe disease at ED disposition (p=0.014) and within 30-days (p<0.001).

**Table 3.** Cytokine and Inflammatory Ratios.

| Ratio (unit) | Admission Severity |  |  | Maximum Severity |  |  |
| --- | --- | --- | --- | --- | --- | --- |
|  | Mild/Moderate<br>(n=46) | Severe<br>(n=6) | p-value | Mild/Moderate<br>(n=36) | Severe<br>(n=16) | p-value |
| **IL-6/IL-10** | 7.0 (5.4-13.7) | 8.5 (2.1-27.1) | 0.827 | 7.4 (5.5-13.4) | 6.9 (4.6-19.8) | 0.734 |
| **IL-10/TNF-α** | 0.32 (0.15-0.88) | 1.3 (0.48-9.5) | 0.021 | 0.3 (0.3-0.8) | 0.6 (0.5-3.3) | 0.020 |
| **IFN-γ/ IL-10** | 83.4 (42.4-157.9) | 25.1 (2.2-25.1) | 0.07 | 92.5 (49.2-141.7) | 43.2 (19.9-148.7) | 0.094 |
| **IL-6/lymphocyte count** | 12.7 (4.8-39.8) | 51.7 (23.7-415.9) | 0.014 | 11.5 (3.9-27.0) | 39.5 (16.5-81.4) | 0.001 |
| **IL-10/lymphocyte count** | 1.6 (0.50-3.1) | 11.4 (7.9-31.0) | 0.008 | 1.1 (0.4-2.6) | 6.4 (1.6-11.6) | <0.001 |

\* All data presented as median (IQR). \*\*IL – interleukin, TNF – Tumor Necrosis Factor

Data on IL-10, IL-10/lymphocyte count, and IL-10/TNF-α with respect to secondary and tertiary outcome is presented in Table 4. IL-10 and IL-10/lymphocyte count were significantly elevated in patients who developed severe AKI during hospitalization and in those with who developed a secondary bacterial infection (all p’s ≤ 0.01). IL-10/TNF-α ratio was not associated with either outcome.

**Table 4.** **Compensatory Anti-inflammatory Response, Acute Kidney Injury, and Secondary Bacterial Infections**

| Variable | No Severe AKI (None or KDIGO 1) | Severe AKI (KDIGO 2+3) | p-value | No Secondary Bacterial Infection | Secondary Bacterial Infection | p-value |
| --- | --- | --- | --- | --- | --- | --- |
| **IL-10** | 1.2 (0.5-2.5) | 5.6 (1.5-8.5) | 0.007 | 1.2 (0.5-2.5) | 5.8 (2.6-8.1) | 0.001 |
| **IL-10/TNF- $\alpha$** | 0.4 (0.2-0.9) | 0.5 (0.1-3.7) | 0.761 | 0.3 (0.2-0.9) | 0.5 (0.3-3.3) | 0.204 |
| **IL-10/lymphocyte count** | 1.6 (0.5-3.0) | 8.4 (1.1-11.8) | 0.010 | 1.6 (0.5-3.0) | 9.2 (3.4-11.6) | 0.003 |

\* All data presented as median (IQR). \*\*IL – interleukin, TNF – Tumor Necrosis Factor, AKI – Acute Kidney Injury

IL-10 and IL-10/lymphocyte count ratio were analyzed in multivariate logistic regression for independent associated with outcomes (Table 5). After adjusting for confounders, a one-unit increase in IL-10 was associated 42% increased odds of severe COVID-19 within 30 days of index ED visit (p=0.031), whilst a one-unit increase IL-10/lymphocyte count ratio was also associated with 32% increased in odds of odds of severe COVID-19 within 30 days of index ED visit (p=0.013). Only IL-10 was found to be predictive of severe AKI in multivariate analysis, with a one-unit increase associated with 40% increased odds of this outcome. Logistic regression for secondary bacterial infections was prohibited due to the low number of patients experiencing this complication.

**Table 5.** **Results of Multivariate Logistic Regression for Severe COVID-19 and Severe Acute Kidney Injury.**

| **Variable** | **Estimate** | **Standard Error** | **p-value** | **OR (95% CI)** |
| --- | --- | --- | --- | --- |
| **IL-10 and Peak 30-Day COVID-19 Severity** |  |  |  |  |
| Age | 0.076 | 0.031 | 0.013 | 1.08 (1.02 - 1.15) |
| IL-10 | 0.354 | 0.164 | 0.031 | 1.42 (1.03 - 1.96) |
| **IL-10/Lymphocyte Count and Peak 30-Day COVID-19 Severity** |  |  |  |  |
| Age | 0.074 | 0.032 | 0.004 | 1.08 (1.01 - 1.15) |
| IL-10/Lymphocyte Count | 0.279 | 0.113 | 0.013 | 1.32 (1.06 - 1.65) |
| **IL-10 and Severe AKI** |  |  |  |  |
| Number of Comorbidities | 0.336 | 0.141 | 0.017 | 1.39 (1.06 - 1.84) |
| IL-10 | 0.337 | 0.160 | 0.035 | 1.40 (1.02 - 1.92) |

## 4. DISCUSSION

The results of this study demonstrate that the pro-inflammatory response to SARS-CoV-2 infection is accompanied by a substantial anti-inflammatory response, which is associated with disease severity even when measured at ED presentation. While the hyperinflammatory consequences of COVID-19 has been the focus of much debate, our study emphasizes that concurrent CARS/immunoparalysis contributes to the “cytokine storm” immunopathology and clinical outcomes of disease. Hence, the notion of COVID-19 as a purely hyperinflammatory condition is likely to be a flawed oversimplification. The relative contribution of hyperinflammation and immunoparalysis would be expected to differ among patients and during the different stages of disease, which complicates treatment strategies and clinical trials using immunomodulatory strategies. Immunomodulation must be tailored to the underlying immunobiology for any given patient, and we support the proposition of Hall et al. that personalized immunophenotype-based immunotherapy is likely to yield superior outcomes [13]. Failure of IL-6 inhibition in clinical trials may have resulted from a heterogeneous group, including patients with significant immunoparalysis and variable immunophenotypes at time of administration.

In this investigation, we observed an independent association between disease severity and IL-10 and IL-10/Lymphocyte Count Ratio. In sepsis, high values of IL-10 are associated with poor outcomes, including higher mortality.[14] In a finding similar to COVID-19, the CARS of severe sepsis is associated with high IL-10 and lymphocyte depletion [10]. Li et al. reported that the IL-10/Lymphocyte Count Ratio was associated with severity and outcome of infection in sepsis patients admitted to the intensive care unit [10]. In COVID-19, the high IL-10 appears to be driven by the overall dysregulated inflammatory response as denoted by the significant correlation with IL-6 observed in this study. Moreover, this cytokine storm likely inhibits the utility of directly comparing pro-versus anti-inflammatory cytokines. As found in this study, though IL-10/TNF-α ratio was significant when comparing levels by disease severity in univariate analysis, this was not confirmed in multivariate analysis. As such, comparing a cytokine with a cellular marker may yield more informative assessment.

It has already been established that low lymphocyte count is associated with poor disease progression and outcomes in COVID-19 [15–17]. The mechanism for the observed low lymphocyte counts in patients with severe COVID-19 is not fully elucidated, but likely involves a combination of disrupted bone marrow generation, cell depletion, apoptosis, and direct viral cytopathic insults [18]. Compounding these, IL-10 may play a part in the apoptosis of lymphocytes, as observed in sepsis [19–21]. Yet both T-cell and B-cell lymphocytes secrete IL-10 [22], thus demonstrating the intricate relationship between these two measures of immune status. The combination of lymphopenia with elevated IL-10 reflects a state of functional immunoparalysis, which would be expected to impair both innate and adaptive antimicrobial responses.

Along with its association with disease severity, elevated IL-10 levels at ED presentation were also found to be independently associated with increased odds of developing severe AKI. At first glance, this may be relatively surprising, as IL-10 is a potent anti-inflammatory cytokine that inhibits the inflammatory and cytotoxic pathways that perpetuate AKI [23]. Animal models have demonstrated that IL-10 attenuates inflammation and AKI due to a variety of insults, including ischemic injury [24]. This is thought to occur for a number of mechanisms, including the inhibition of genes triggering leukocyte activation and adhesion, and inducing inducible nitric oxide synthase [24]. However, Zhang et al. reported that IL-10 elevation was associated with AKI (adjusted OR: 1.57 [95% CI, 1.04 to 2.38]) but lower risk of mortality (adjusted hazard ratio, 0.72 [95% CI, 0.56 to 0.93]) in adults after cardiac surgery [25]. In part, IL-10 exerts its protective renal effects through induction of neutrophil gelatinase-associated lipocalin (NGAL) [26], which itself is an early and sensitive diagnostic and prognostic serum and urine biomarker of AKI, which can rise 24 hours earlier than creatinine [27]. In further analysis, Zhang et al. observed that improved outcomes in patients with high IL-10 levels could only be seen in those with concomitant elevations in NGAL, while those with high IL-10 and low NGAL were afforded a worse prognosis [25]. Such findings suggest that IL-10 may only be protective when it induces NGAL expression [25]. During the cytokine storm of COVID-19, in which a dysregulated immune response occurs, we suspect that there may be minimal expression of NGAL, thus explaining the findings observed with respect to IL-10 in this study. Therefore, NGAL shall be measured and correlated with IL-10 in future studies on patients with COVID-19 to confirm this hypothesis, as well as for assessing overt AKI and subclinical renal injury in patients with COVID-19.

Unsurprisingly, IL-10 and IL-10/Lymphocyte Count were significantly elevated in patients who developed secondary bacterial infections. This finding suggests a direct clinical consequence of functional immunoparalysis in some patients with severe COVID-19 driven by high IL-10 blunting a pro-inflammatory antimicrobial response with low lymphocyte count impairing adaptive immunity. In general, secondary infections are associated with increased morbidity and mortality in viral respiratory infections, including COVID-19 [28–30]. In a comprehensive meta-analysis, Langford et al. reported a prevalence rate of 14.3% (95%CI: 9.6-18.9%) for secondary bacterial infections in patients with COVID-19 [31]. However, the authors argued against empiric antibacterial therapy [31]. We suggest antibiotic therapy may be tailored according to immunophenotype, with elevated IL-10/Lymphocyte Count ratio serving as a threshold marker for initiation of empiric antibiotic therapy. Due to the limited number of subjects with secondary infections in our study, we could not evaluate this variable in multivariate analysis. However, this should be further investigated in future studies.

This study was limited by a relatively small sample size and a single time point of laboratory measurements. We choose ED presentation as a time point of clinical significance, as it is not only the point when a patient felt sick enough to seek medical care, but it often presents one of the earliest and perhaps most efficient opportunity for intervention. Although patients’ immunologic state may significantly vary over the course of illness, we were able to capture patients at a variety of stages at index ED visit, including severe disease. Nonetheless, future studies should be performed to serially monitor biomarkers of immunoparalysis throughout hospitalization. Moreover, future molecular studies are needed to characterize the CARS response, including HLA-DR monocyte expression and ex vivo LPS-induced TNF-α release. Finally, when pre-COVID-19 serum creatinine value was not available, the index ED visit serum creatinine was used in such cases as a baseline value for evaluating AKI. Thus, it is possible that cases of AKI were missed in patients who presented with peak creatinine value resolving during hospitalization. As such, the incidence of this endpoint may have been underrepresented.

## 5. CONCLUSION

The hyperinflammatory response to COVID-19 is accompanied by a simultaneous CARS that is associated with poor outcomes and may increase the risk of secondary bacterial infections. IL-10 and IL-10/lymphocyte count ratio at ED presentation were independent predictors of COVID-19 severity, while IL-10 levels predicted development of AKI. The CARS phenomenon further introduces heterogeneity into COVID-19 population, and may mask effects of potentially useful anti-inflammatory drugs for patients without substantial CARS in clinical trials. The CARS during COVID-19 requires further investigation to enable more precise immunomodulatory therapy against SARS-CoV-2, which should be optimally tailored to an individual patient’s immunophenotype.

## Data Availability

Data available upon reasonable request.

## Funding

This study was funded by the University of Cincinnati College of Medicine Special Coronavirus (COVID-19) Research Pilot Grant Program.

## Conflicts of Interest

The authors declare no conflict of interest.

## Notes

### Competing Interest Statement

The authors have declared no competing interest.

### Author Declarations

This study was approved by the Institutional Review Board of the University of Cincinnati and performed under a waiver of informed consent. This study was performed in agreement with the Declaration of Helsinki and in compliance with local and national regulations.

